# Post-COVID syndrome is associated with capillary alterations, macrophage infiltration and distinct transcriptomic signatures in skeletal muscles

**DOI:** 10.1101/2023.02.15.23285584

**Authors:** Tom Aschman, Emanuel Wyler, Oliver Baum, Andreas Hentschel, Franziska Legler, Corinna Preusse, Lil Meyer-Arndt, Ivana Büttnerova, Alexandra Förster, Derya Cengiz, Luiz Gustavo Teixeira Alves, Julia Schneider, Claudia Kedor, Rebecca Rust, Judith Bellmann-Strobl, Sanchin Aminaa, Peter Vajkoczy, Hans-Hilmar Goebel, Markus Landthaler, Victor Corman, Andreas Roos, Frank L. Heppner, Helena Radbruch, Friedemann Paul, Carmen Scheibenbogen, Werner Stenzel, Nora F. Dengler

## Abstract

The SARS-CoV-2 pandemic not only resulted in millions of acute infections worldwide, but also caused innumerable cases of post-infectious syndromes, colloquially referred to as “long COVID”. Due to the heterogeneous nature of symptoms and scarcity of available tissue samples, little is known about the underlying mechanisms. We present an in-depth analysis of skeletal muscle biopsies obtained from eleven patients suffering from enduring fatigue and post-exertional malaise after an infection with SARS-CoV-2. Compared to two independent historical control cohorts, patients with post-COVID exertion intolerance had fewer capillaries, thicker capillary basement membranes and increased numbers of CD169^+^ macrophages. SARS-CoV-2 RNA could not be detected in the muscle tissues, but transcriptomic analysis revealed distinct gene signatures compared to the two control cohorts, indicating immune dysregulations and altered metabolic pathways. We hypothesize that the initial viral infection may have caused immune-mediated structural changes of the microvasculature, potentially explaining the exercise-dependent fatigue and muscle pain.

## INTRODUCTION

Persisting general or muscular fatigue are well-known phenomena occurring in a proportion of previously healthy individuals after an acute viral infection, and many endemic or epidemic outbreaks of such symptoms have been documented (*1-5*). While most people infected with SARS-CoV-2 recover without sequelae, subsets of both outpatient and hospitalized individuals experience newly occurring and lasting symptoms such as fatigue, muscular weakness, exercise intolerance with post-exertional malaise (PEM), myalgia or neurocognitive and neuropsychiatric disturbances (*6-11*). The pathomechanisms behind *post-COVID Syndrome* (PCS), *Post-acute sequelae of SARS-CoV-2 infection* or *long COVID*, are not well understood, plausibly because the manifestations and causes are diverse and semantically diffuse, thus hampering comparative descriptive and mechanistic studies. Similar to post-SARS-syndrome reported after the SARS epidemic in 2003-2004 (*12, 13*), many clinicians noticed that some prevailing symptoms of PCS were reminiscent of chronic fatigue syndrome (ME/CFS) (*14-17*). This syndrome is entirely based on clinical criteria and exclusion of other diagnoses, plausibly consisting of different causal entities resulting in a similar phenotype, which is centered around a chronic fatigue aggravated by physical and/or mental activity and accompanied by other symptoms (e.g. sleep disturbances, neurocognitive deficits), leading to significant restrictions in everyday life (*17-21*). In the majority of patients, ME/CFS is triggered by an infection (*22-28*).

While inflammatory myopathy has been shown in individuals with fatal COVID-19 (*29, 30*), to our knowledge no case-control studies investigated the affection of skeletal muscle tissue in individuals with PCS. We therefore designed this case-control study of vastus lateralis muscle biopsies obtained from individuals suffering from chronic muscular fatigue, myalgia and PEM after a PCR-proven infection with SARS-CoV-2. Biopsies were taken in average one year after initial infection and were compared to historical control samples, consisting of histologically normal samples as well as samples with a selective atrophy of type-2b muscle fibers. The latter is a non-specific finding associated with muscle disuse and immobility. While patients with PCS did not show overt signs of myositis, increased numbers of CD169^+^ macrophages were evident. The capillary-to-fiber ratio was decreased and ultrastructural analysis revealed a capillary basement membrane enlargement. SARS-CoV-2-specific RNA was not detected, but transcriptomic analysis showed distinct transcriptomic profiles in most of the PCS samples compared to the controls, with increased expressions of genes related to immune system regulation, angiogenesis and extracellular matrix remodeling, and decreased expressions of genes related to metabolic processes and mitochondrial activity.

## RESULTS

### Clinical characteristics of the post-COVID syndrome cohort

Eleven individuals (female-to-male ratio 10:1; mean age 45 years, [SD 11.5]; range 25 – 58 years) who were tested positive for SARS-CoV-2 by PCR between March 2020 and December 2020 and who subsequently developed a new fatigue with muscular symptoms (weakness, myalgia) and exertion intolerance with post-exertional malaise (PEM) lasting for at least 6 months which could not be explained by alternative diagnoses underwent a biopsy of vastus lateralis muscle, fascia and skin. Nine of these patients were part of a larger observational study during which they were asked to submit subjective symptom surveys at defined intervals (*14*). The majority of patients (91%; n=10/11) only had mild COVID as defined by the WHO (*31*). One patient however (*PCS-11*) was hospitalized and required four days of non-invasive ventilation due to dyspnea, thus reaching a score of 6 (severe COVID) (*31*). The timeline of events, as well as the documented symptoms of acute and chronic are reported in Fig. 1A-B and Table S1.

**Fig. 1.**
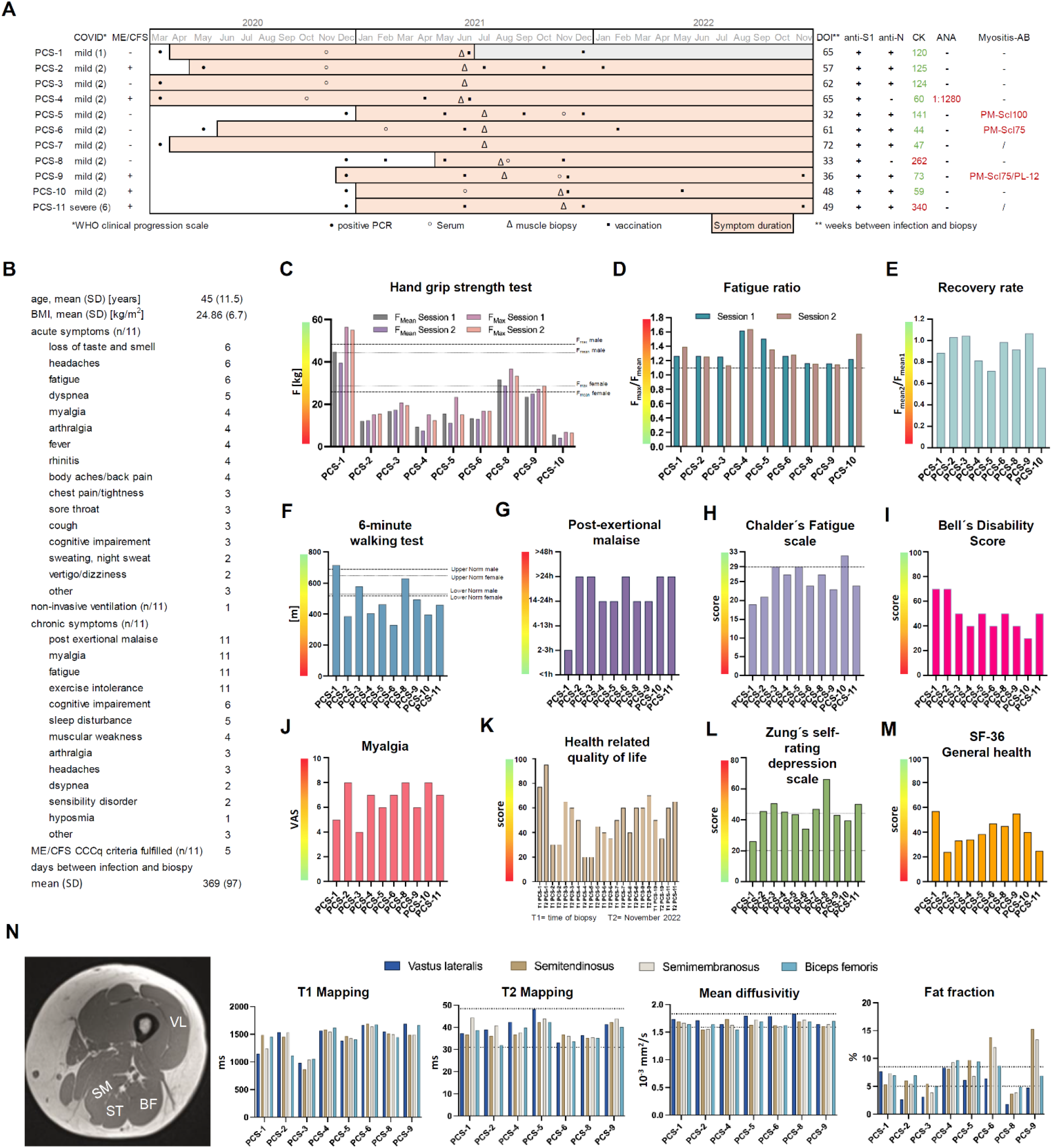
Clinical characteristics of the post-COVID syndrome cohort. (**A**) Demographics of study participants with Post-COVID Syndrome (PCS) and timeline of events. Black dots indicate the month in which PCR-testing for SARS-CoV-2 was positive. Black circles indicate the month from which serum samples were available. For two patients (*PCS-7* and *PCS-11*), no serum was available for mass spectrometry and myositis-specific and –associated antibody screening. Black triangles indicate the month in which biopsy of vastus lateralis muscle was performed. Black cubes indicate the months of anti-SARS-CoV-2 vaccinations. Colored bars indicate the months in which the participants reported symptoms. (**B**) Patients characteristics, summary of acute and chronic symptoms. (**C**) Handgrip strength test (HGS) was performed in two subsequent sessions (session 1 and 2) in 9/11 patients around the time of biopsy. Dotted lines indicate the mean reference values based on ROC analyses showing the highest sensitivity and specifity to discriminate between healthy controls and ME/CFS patients for men and women respectively. (**D**) Fatigue ratio defined as the ratio of F_max_ / F_mean_ per individual session. Dotted lines indicate the respective normal population mean reference values for women and men. (**E**) Recovery rate defined as the ratio of F _mean_ of session 2 divided by F _mean_ of session 1. Dotted lines indicate the normal population mean reference value. (**F**) Standardized six-minute-walk test was performed in 10/11 individuals. Dotted lines indicate the upper and lower reference values for men and women respectively. (**G**) Duration of post-exertional malaise around the time of biopsy. (**H-M**) Subjective scores around the time of biopsy. Dotted lines indicate reference values, when present for the German population. (**N**) MRI of proximal lower extremity with indication of ROI (regions of interest) and results of T1 and T2 mapping, mean diffusivity and fat fraction. *Abbreviations: HDC= healthy-diseased control; PCS= post-COVID syndrome; 2BA= type-2b atrophy control; BMI=body mass index; SD= standard deviation; ME/CFS= Myalgic encephalomyelitis/chronic fatigue syndrome; DOI= duration of illness (at timepoint of biopsy); CK= creatine kinase; ANA= anti-nuclear antibodies; Myositis-AB= myositis-associated or - specific antibodies); F= force*.

All patients complained of persisting fatigue with exercise intolerance and myalgia at the time of study inclusion. In all but one patient, symptoms were still present by the end of November 2022 and five patients fulfilled the Canadian Consensus Criteria (CCC) for ME/CFS (*32*). The average time between onset of acute infection symptoms or positive PCR and muscle biopsy was 369 days (SD 97) (Fig. 1A-B).

Standardized handgrip strength test (HGS) was performed in nine, and a 6-minute-walk-test (6MWT) in ten patients. Compared to reference values (*33*), seven patients (78%; n=7/9) showed abnormal values for maximal and mean force, fatigue ratio and recovery rate in the HGS (Fig. 1C-E). Seven patients (70%; n=7/10) walked a shorter distance within six minutes than expected for their age and sex(*34*) (Fig. 1F). Most patients reported a PEM of at least 14 hours (Fig. 1G). Perceived symptoms and impairments were assessed by standardized questionnaires, including the *Bell disability score*^*i*^ (*0-100; 0= bedridden, 100= healthy), Chalder fatigue score (35), SF-36 (36), EQ-5D-5L (37, 38*) and *Zung Self-Rating Depression Scale (39*). Ten individuals (91%) scored 50 or lower on the Bell disability score, indicating a functionality level below 70% compared to the German reference population (Fig. 1I). The mean individual health related quality of life on the visual analog scale from 0-100 in our patient cohort was 50.8 (SD 18.7), being substantially lower than the average German population which was shown to be 79.5 (SD 17.1) (Fig. 1K) (*38*). In our patient cohort, the EQ-5D-5L mean scores for the dimensions mobility, self-care, usual activities, pain and anxiety on a scale from 1 (no problems) to 5 (extreme problems) were 3/2/4/4/2, indicating moderate affection in the dimensions mobility and severe problems within the dimensions usual activities and pain, respectively. Most of these dimension’s scores as well as other functional scores did not evolve for the better in some patients (*PCS-2, PCS-3, PCS-4, PCS-6, PCS-10*) while they improved to some degree in others (*PCS-1, PCS-5, PCS-7, PCS-8, PCS-9, PCS-11*) (Fig. 1K & Fig. S1A).

Proximal lower extremity MRI was performed in nine patients on the same day or close to the day of biopsy. This revealed no radiological evidence for myositis in the biceps femoris (BF), semitendinosus (ST), semimembranosus (SM), and vastus lateralis (VL) muscles. T2 mapping and mean diffusivity were comparable to levels of healthy control cohorts from published studies (*40-43*). Muscle Fat Fraction (MFF), depending on age, physical fitness, sex and hormonal components were comparable to reference values in most subjects, but two study participants (*PCS-6* and *PCS-9*) showed slightly increased values in two of the four examined muscles (*44, 45*) (Fig. 1N). Comparability to published data is however limited due to technical differences in acquisition.

Heart MRI revealed a discrete pericarditis in one patient (*PCS-4*), that had disappeared in a follow-up examination 8 months later. Diffuse fibrotic changes in one other patient (*PCS-5*) and a dilated cardiomyopathy in yet another patient (*PCS-11*) were noted (Table S1). Levels of creatine kinase were below the lab threshold (<167 U/L) in all but two patients, who only had slightly increased levels (*PCS-8*: 1.5-fold increase; *PCS-11*: 2-fold increase), and one patient (*PCS-4*) had high titers of antinuclear antibodies (1:1280) with a dense fine speckled nuclear staining pattern, without detection of specific autoantibodies (Fig. 1A). Screening for myositis-specific and myositis-associated autoantibodies revealed the presence of anti-PM-Scl75 antibodies in two patients, one of them also having anti-PL-12 antibodies, and anti-PM-Scl100 antibodies in another patient (Fig. S2).

### Virology

All patients from the PCS cohort were tested positive for SARS-CoV-2 by PCR from nasopharyngeal swabs in the course of 2020. Presence and titers of antibodies against SARS-CoV-2 spike (S) and nucleocapsid (N) antigen were determined using two different techniques (ELISA and *ECLIA*). This revealed a presence of anti-S-IgG in all patients and of anti-N-IgG in 9/11 patients. One of the two patients without anti-N-IgG had detectable anti-S-IgG above the positivity threshold already prior to vaccination (Fig. 1A), also proving a contact with SARS-CoV-2 (Fig. S1B).

Quantitative reverse transcription–polymerase chain reaction (RT-qPCR) of skeletal muscle specimen did not yield SARS-CoV-2 specific RNA amplification below the lab threshold cycle. In one sample (*PCS-4*), a first reaction gave a weakly positive result that could not be confirmed in a repeat experiment.

### Type-2b-fiber atrophy and increased numbers of tissue macrophages in skeletal muscles of patients with PCS

Routine histopathological examination revealed a selective atrophy of type-2b muscle fibers of differing extent in 72% (n=8/11) of individuals with PCS and – by definition of our inclusion criteria – in none (n=0/8) of the HDC cohort and in all (n=8/8) of the 2BA cohort (Fig. 2A). In one patient of the PCS cohort (n=1/11) one single necrotic fiber was observed, whereas none were found in the HDC and 2BA cohort. No sarcolemmal expression of C5b-9 was found in any sample, but in one patient from the PCS cohort (*PCS-11*) a weak capillary expression of C5b-9 was present (Fig. S3B). Single muscle fibers with sarcolemmal upregulation of MHC-class-I were found in two samples from the HDC cohort, in one from the 2BA and in three from the PCS cohort. These were judged as non-pathological. Two samples from the PCS cohort (*PCS-3* and *-10*) but none from the two control cohorts revealed a significant upregulation of MHC-class-I of at least 20% of the muscle fibers (Fig. 2C). In one of these patients (*PCS-3*) a sarcolemmal upregulation of MHC-class-II was also present on some fibers (Fig. S3C). Upregulation of Interferon-induced GTP-binding protein Mx1 was only observed on several capillaries in one sample from the PCS cohort (*PCS-6*), but in none of the controls (Fig. S3D).

**Fig. 2.**
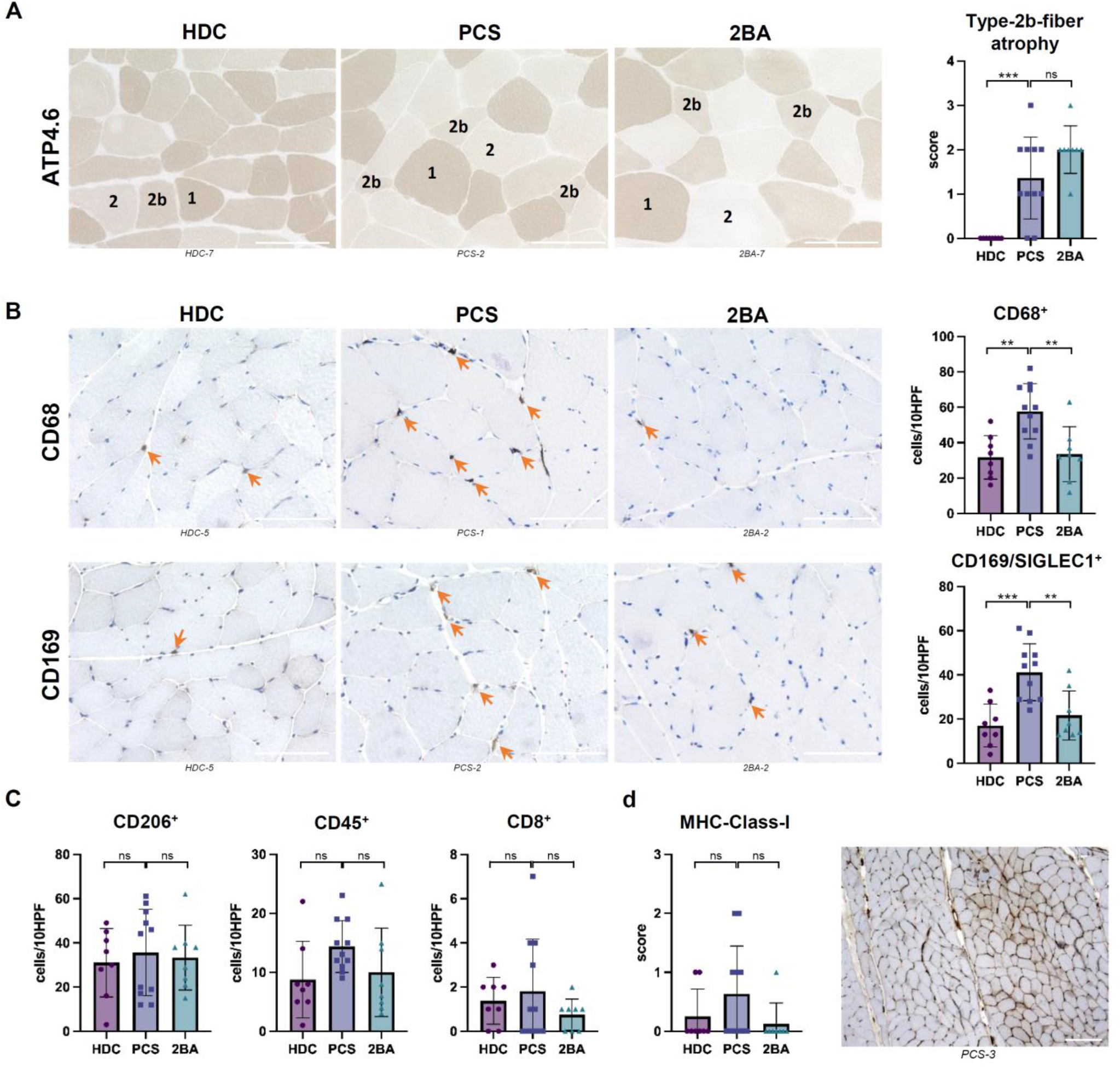
Type-2b-fiber atrophy and increased numbers of tissue macrophages in skeletal muscles of patients with PCS. (**A**) ATP4.6 enzymatic reaction showing selective atrophy of type-2b muscle fibers in PCS samples and the 2BA control cohort. (**B**) Immunohistochemistry and quantification of CD68^+^ and CD169^+^ macrophages. (**C**) Immunohistochemistry and quantification of CD206^+^, CD45^+^ and CD8^+^ cells. (**A-C**) 400x magnification, scale bar = 100µm. (**D**) Immunohistochemistry, semi-quantitative scoring of sarcolemmal MHC-cl.-I expression and example of a sample with a score of 2 (*PCS-3*). 100x magnification, scale bar= 200µm. Ordinary one-way ANOVA with Tukey’s multiple comparisons test. * = p<0.05, ** = p<0.005, *** = p <0.0005. *Abbreviations: ATP4*.*6= Adenosine 5’-TriPhosphatase at pH 4*.*6; SIGLEC1= Sialoadhesin; MHC cl. I= major histocompatibility complex class I*.

Quantification of immune cells revealed a significant increase of CD68^+^ (*HDC vs. PCS*: mean of 32 [SD 12] versus 58 [SD 16] cells/10HPF, p=0.002; *2BA vs. PCS*: mean of 34 [SD 15] versus 58 [SD 16] cells/10HPF, p=0.005) and CD169^+^ (*HDC vs. PCS*: mean of 17 [SD 10] versus 41 [SD 13] cells/10HPF, p= 0.0004; *2BA vs. PCS*: mean of 22 [SD 11] versus 41 [SD 13] cells/10HPF, p= macrophages. Numbers of CD45^+^ cells, CD8^+^ T-cells and CD206^+^ macrophages were not significantly different between the three groups (Fig. 2B-C).

### Decreased capillary-to-fiber ratio and increased capillary basement membrane thickness in patients with PCS

Numbers of capillaries per muscle fiber (C/F) were quantified by assessing at least 10 images of semithin cross-sections of all included muscle specimens at 200x magnification. This revealed a significant decrease in the PCS cohort when compared to the two historical control cohorts (*PCS vs. HDC*: mean difference 0.32, p=0.007; *PCS vs. 2BA*: mean difference 0.30, p=0.013). Comparison of mean cross-sectional fiber area (MCSFA) revealed a significant decrease in the PCS cohort compared to the 2BA cohort (mean difference 1508 µm^2^, p=0.009) whereas no significant difference was present when compared to the 2BA cohort (Fig. 3A). The reduced MCSFA explains why there was no significant difference of the capillary density between the groups. In fact, capillary density represents the absolute number of capillaries per mm^2^ and due to a decreased MCSFA, more fibers and more capillaries will be observed in one field of vision. Therefore, interpretation of the C/F is more conclusive.

**Fig. 3.**
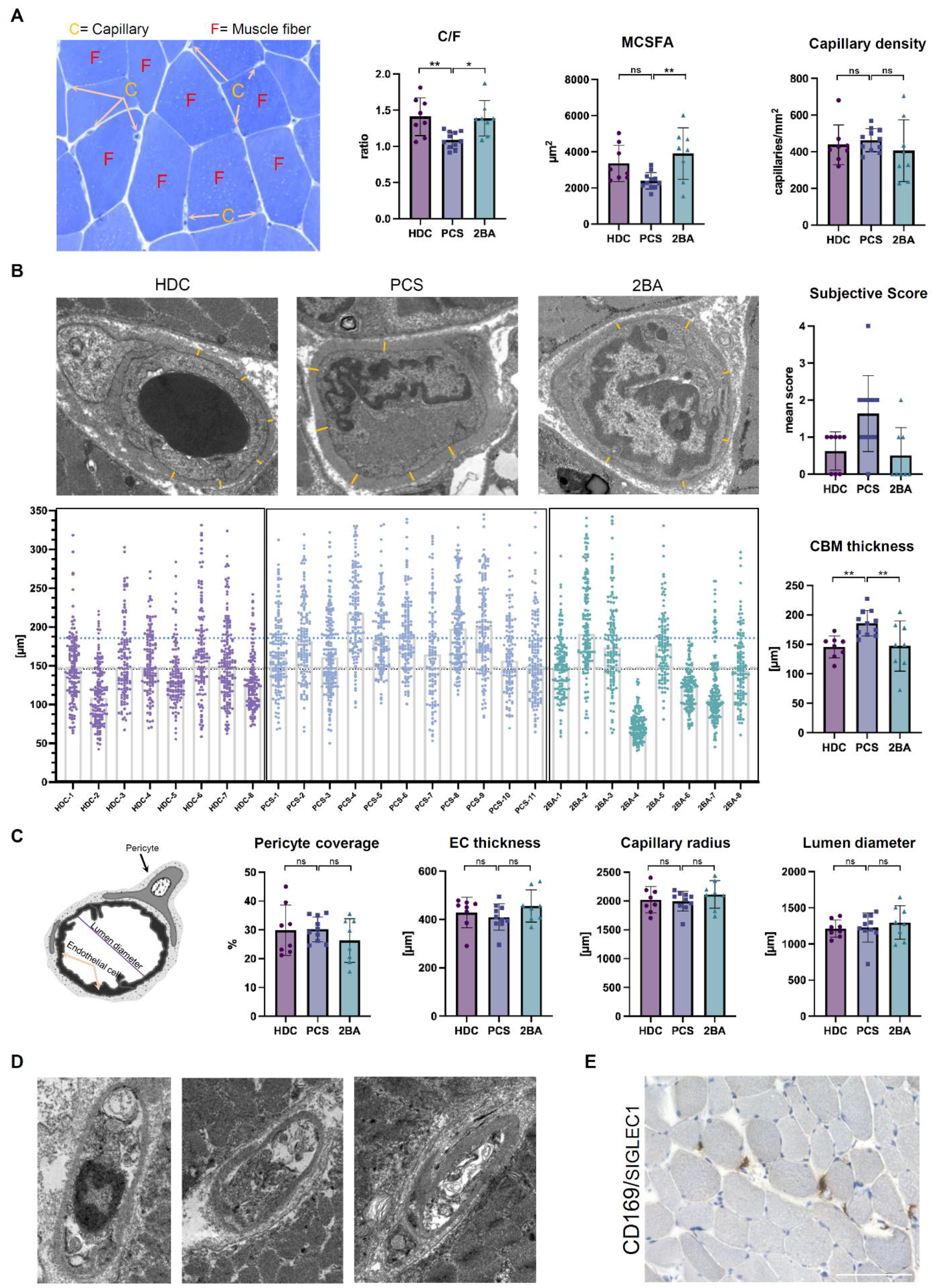
Decreased capillary-to-fiber ratio and thicker capillary basement membranes in patients with PCS. (**A**) Example of a semithin transverse section of a vastus lateralis muscle biopsy specimen stained with toluidine blue, displaying capillaries and muscle fibers, and quantification of C/F, MCSFA and capillary density. (**B**) Representative electron micrographs of transverse ultra-thin sections of muscle capillaries. Orange lines indicate typical sites of measurements for the capillary basement membrane thickness. The large graphs depicts all 3732 individuals measurements except 35 outliers with values above 350µm. Small graphs indicate subjective scoring of capillary pathology as well as mean CBM thickness values per individual samples. (**C**) Schematic cross-sectional representation of a capillary with results of TBIA morphometric analysis. (**D**) Exemplary micrographs of transverse ultra-thin sections of damaged muscular capillaries from *PCS-4*. (**E**) Immunohistochemical expression of CD169/SIGLEC1 in the biopsy specimen obtained from *PCS-4*. 400x magnification, scale bar = 100µm. Ordinary one-way ANOVA with Tukey’s multiple comparisons test. * = p<0.05, ** = p<0.005, *** = p <0,0005. *Abbreviations: C/F= capillary-to-fiber ratio; MCSFA = mean cross-sectional fiber area; CBM= capillary basement membrane; EC= endothelial cell; TBIA= Tablet-based image analysis*

Between 20 and 30 capillaries per sample were photographed by TEM at a magnification of 7000x (mean capillaries per group: *HDC* 23.63 (SD 4), *2BA* 22.75 (SD 2) and *PCS* 24.27 (SD 3). For each individual capillary, the thickness of capillary basement membrane (CBM) was measured at six different sites, excluding areas with profiles of pericyte processes as the variation of CBM is more pronounced there (*46*). This resulted in a total of 3732 single CBM thickness values (Fig. 2B). CBM thickness was significantly increased in the PCS cohort (*PCS vs. HDC*: mean difference 39.99µm, p=0.016; *PCS vs. 2BA*: mean difference 38.48, p=0.021). When excluding the apparent outlier from the 2BA cohort (*2BA-4*) the threshold for significance in that comparison was just not reached (*PCS vs. 2BA*: mean difference 28.54µm, p=0.06).

Morphometry by TBIA revealed no significantly different values for lumen diameters and capillary radius between the groups. Furthermore, no difference regarding pericyte coverage and endothelial cell thickness were observed (Fig. 3C).

In one patient (*PCS-4*), massive structural damage was observed in all 23 photographed capillary profiles. Endothelial cells were almost completely degenerated, resulting in debris-containing empty capillary tubes also called string vessels (=acellular capillary remnants) (Fig. 3D). Such severe morphological alterations were not found in any other patient. Immunohistochemistry revealed the presence of many large CD169^+^ macrophages in close vicinity to the capillaries (Fig. 3E).

There was a moderate correlation between the thickness of CBMs and the number of CD169^+^ macrophages (Pearson’s r 0.53), the EQ-5D-5L mobility score (Pearson’s r 0.78) and the EQ-5D-5L usual activities score around the timepoint of biopsy (Pearson’s r 0.56), but no correlation with age. There was also a correlation between the mean cross sectional fiber size area (MCSFA) and the capillary-to-fiber ratio (Pearson’s r 0.63) (Fig. S5).

### Increased expression of basement membrane proteins in muscles from patients with PCS

Mass spectrometric analysis comparing muscle specimen of patients with PCS to the HDC cohort revealed significantly higher levels of mostly matrisome proteins related to or constituting basement membranes (COL4A2, NID1, HSPG2, LAMC1, LAMA2, LAMB2, FLNA) or muscle fibers (MYH2, SGCB), albeit below the threshold of 2-fold change. Of interest, most of these proteins also had significantly higher expression levels when comparing samples from patients with PCS to samples from our 2BA cohort (Fig. 4A, C-D). This matches our observation described above that the mean values of CBM thickness in that cohort were closer to the PCS cohort than to the HDC cohort. Other notable proteins with increased expression levels in the PCS cohort are TAGLN2 and CALR. Four proteins were significantly decreased below a threshold of 0.5 (HBG1, IGHV3-74, RCN1 and EIF4B). Unbiased principal component analysis resulted in a clear separation of the HDC from the 2BA and PCS samples, while a less clear separation of the PCS patients from the 2BA samples could be observed (Fig. 4B). Quantitative qRT-PCR did not reveal statistically significant differences in the gene expressions of CBM key components except for NID1 (Fig. 4E). GO-Enrichment analysis revealed an upregulation of biological activity terms related to regulation of basement membrane and extracellular matrix (ECM) organization, as well as cell-cell and cell-ECM signaling (Fig. 4H).

**Fig. 4.**
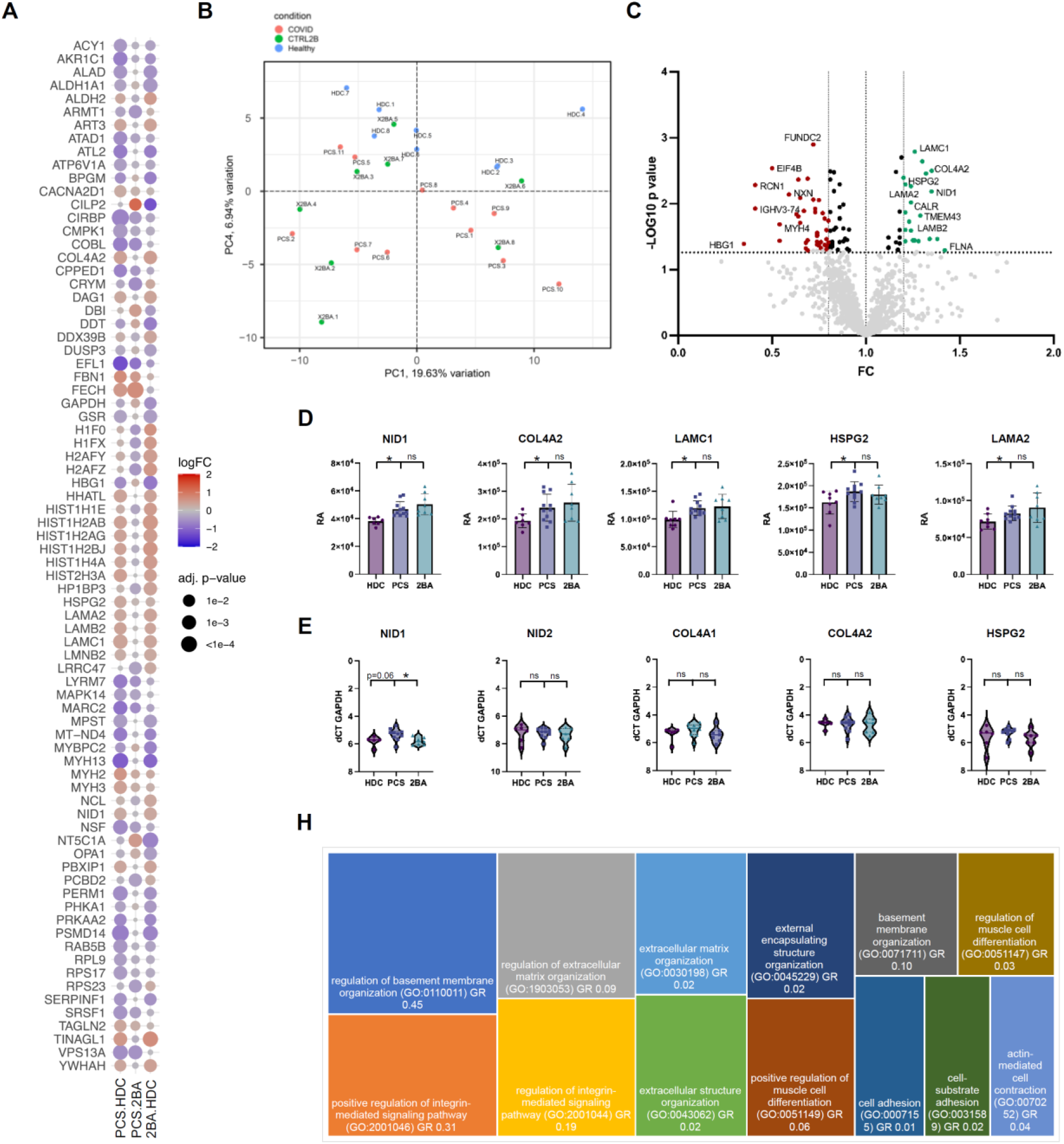
Increased expression of basement membrane proteins in muscles from patients with PCS. (**A**) Dot plot visualization of top differentially abundant proteins comparing all samples from the PCS cohort to the HDC and 2BA cohorts respectively. Dot size is proportional to the adjusted p-value. Dot colour indicates logFC. (**B**) Principal component analysis of proteome raw data after normalization. (**C**) Volcano plot of the vastus lateralis proteome comparison illustrating significantly differentially abundant proteins between the PCS and the HDC cohort. The -log10 (p value) is plotted against the fold change of PCS/HDC. The dotted vertical lines denote +1.2- and -0.8-fold change respectively, while the dotted horizontal line denotes *p* = 0.05. (**D**) Relative abundance (RA) of selected proteins constituting the CBM. (**E**) Quantitative qRT-PCR expression levels of selected genes related to the CBM. (**F**) Treemap visualizing the GO-enrichment analysis of significantly increased proteins. Size of rectangles correlates with -log 10 (p-value). GR= Gene Ratio (Number of Proteins in the indicated GO-term increased in our dataset divided by the total number of proteins included in that GO-term biological activity). Ordinary one-way ANOVA with Tukey’s multiple comparisons test. * = p<0.05.

### Distinct transcriptomic profiles of vastus lateralis muscles from patients with PCS

Bulk RNA sequencing was performed on vastus lateralis muscle specimens obtained from all included individuals. Principal component analysis resulted in a similar clustering by condition as observed in the above-mentioned muscle proteomics analysis. Among the top differentially expressed genes (p≤0.001) were genes related to extracellular matrix remodeling (*ADAMTS4, MMP3*), angiogenesis (*ANGPTL7*) and immune system regulation (*TNF*) (Fig. 5A). Although we observed enriched genes located on the Y-chromosome, corresponding to the sex-misbalance between the HDC and the two other cohorts, we did not see any sex-related influence on the PCA. Similarly, no age-related effect on the PCA was apparent (Fig. 5B-G & Fig. S4A). When excluding all Y-Chromosome related genes, 135 genes were differentially expressed in the comparison PCS vs. HDC (p-value ≤0.001, 114 upregulated, 21 downregulated), 219 genes in the comparison PCS vs. 2BA (p-value ≤0.001, 218 upregulated, 11 downregulated) and 28 in the comparison 2BA vs HDC (p-value ≤0.001, 18 upregulated, 10 downregulated).

**Fig. 5.**
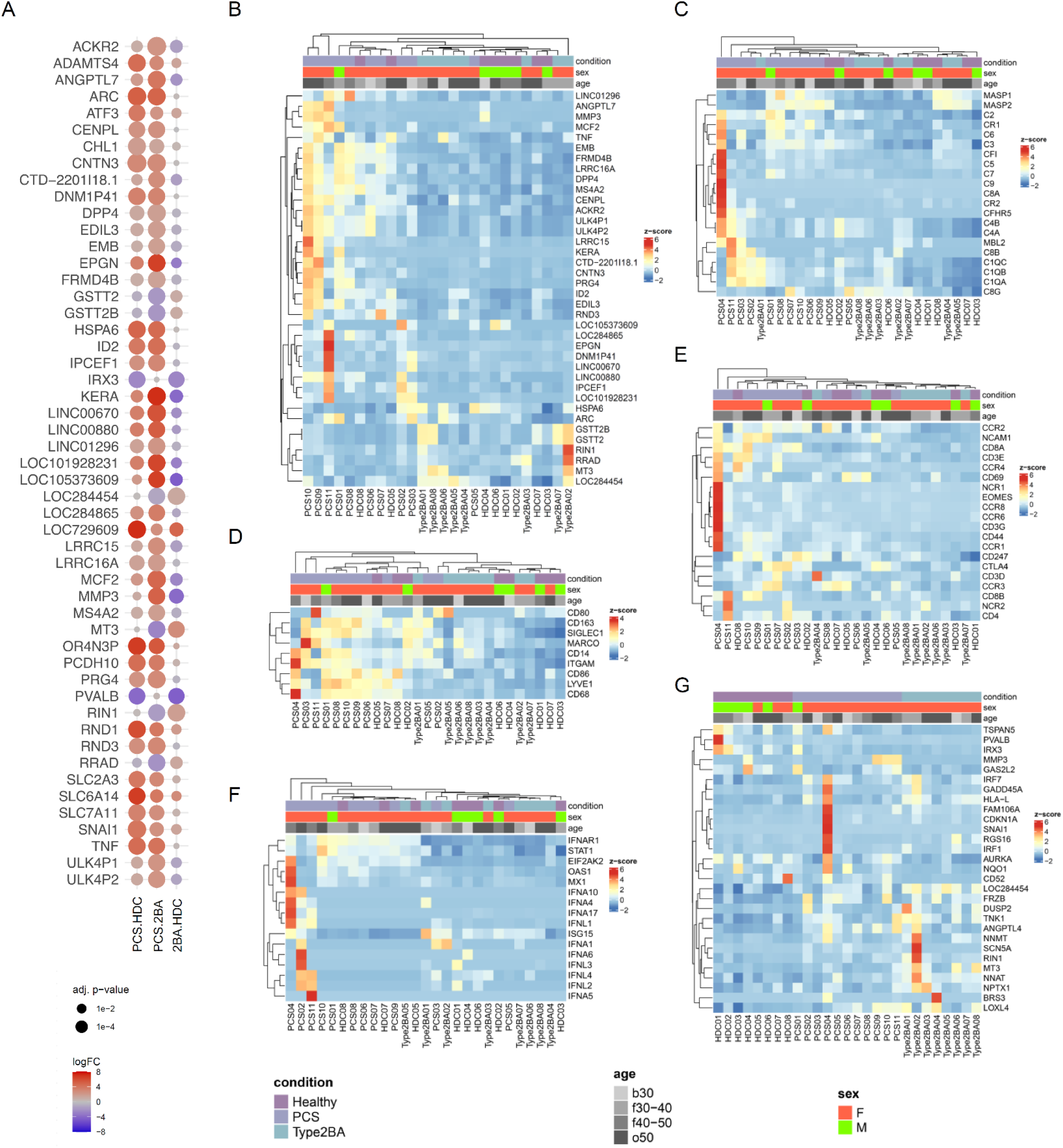
Distinct transcriptomic profiles of vastus lateralis muscles from patients with PCS. (**A**) Dot plot visualization of top differentially expressed genes comparing all samples from the PCS cohort to the HDC and 2BA cohorts respectively. Dot size is proportional to the adjusted p-value. Dot color indicates logFC. (**B**) Clustered heat map visualization of top differentially expressed genes comparing the PCS cohort without *PCS-4* to the HDC and 2BA cohorts. (**C-G**) Clustered heat map visualizations of the gene expressions related to the complement pathway (**C**), of macrophage marker genes (**D**), of diverse immune cell marker genes (**E**) and type I and III interferon related genes (**F**). (**G**) Unclustered heat map visualization of top differentially expressed genes comparing the HDC to the 2BA cohort.

Of note, the patient in which we identified massive structural capillary damage (*PCS-4*) constituted a clear outlier with much higher expression levels of mostly immune system related genes (Fig. 5C-G, & Fig. S4A, C-D).

Unbiased clustering revealed a certain heterogeneity within the PCS group, with some samples showing gene expression levels resembling those of control samples from patients with type-2b-fiber atrophy, while other samples from the PCS cohort showed clearly distinct transcriptomic signatures when compared both to the HDC and 2BA control cohort (Fig. 5B, G).

When clustering all samples by their expression levels for complement pathway or type I and III interferon related genes, many samples from patients with PCS grouped close together and showed a distinct gene signature compared to the two control cohorts (Fig. 5C, E-F). Macrophage marker genes (*CD68, SIGLEC1, CD163, LYVE1, CD86, ITGAM*) were upregulated in the PCS cohort, confirming the histological observations, albeit not reaching a p-value ≤ 0.001 except for CD68 in the comparison PCS vs 2BA (LogFC 0.95, p=0.001) (Fig. 5D). Immune cell markers for T-cells and innate lymphoid cells (ILCs) also lead to co-clustering of the PCS samples (Fig. 5E). Only few differentially expressed genes were found in the comparison of the HDC with the 2BA control cohort (Fig. 5G).

For pathway analysis, all differentially expressed genes with a p-value ≤0.01 were used. Among the top biologically relevant upregulated pathways were “*regulation of leukocyte activation*”, “*positive regulation of cell motility*”, “*vasculature development*” for the comparison PCS vs HDC and “*positive regulation of locomotion*”, “*cell-cell adhesion*”, “*inflammatory response*”, “*NABA Matrisome associated*”, “*chemotaxis*”, “*blood vessel development*”, “*extracellular matrix organization*” and “*NABA Proteoglycans*” in the comparison PCS vs 2BA. Among the top biologically relevant, downregulated pathways were “*mitochondrion organization*”, “*oxidative phosphorylation*” and other mitochondria-related pathways in the comparison PCS vs HDC and “*regulation of carbohydrate biosynthetic process*” in the comparison PCS vs 2BA (Fig. S5E).

## DISCUSSION

The individual and socioeconomic impact of post-COVID syndrome and other post-infectious syndromes such as ME/CSF is considerable (*21, 47, 48*). Affected patients often face a double challenge, the one of the direct physical and mental suffering, and the one of the psychological burden of being affected by a disease with no clear biomarkers and absence of clear-cut, easily observable structural alterations. Therefore, studies on the tissue level as well as translational integrative studies combining clinical observations with histopathological and molecular findings are urgently needed. With all the tragedies related to the SARS-CoV-2 pandemic, it also offers the unique opportunity for studying post-viral syndromes in a more homogeneous manner than previously.

To our knowledge, this is the first case-control study examining skeletal muscle tissue obtained from patients with persisting post-infectious fatigue and exercise intolerance that newly occurred after an infection with SARS-CoV-2. In one descriptive case series lacking controls, histological changes and capillary alterations were described in deltoid muscles of patients with post-COVID syndrome (*49*).

In the present interdisciplinary case-control cohort study, comparing patients with post-COVID syndrome to two distinct age-matched historical control cohorts, we found, on the morphological level, capillary alterations consisting of a decreased capillary-to-fiber ratio and an increased capillary basement membrane thickness. Patients with PCS had smaller muscle fibers and increased numbers of CD169^+^ macrophages in close vicinity to skeletal muscle capillaries, but no evidence of overt myositis. Upper leg MRI also did not reveal signs of myositis, which is consistent with recently published radiological findings (*50*). However, biopsies were taken almost a year after acute infection and several case reports of biopsy-proven myositis after SARS-CoV-2 have been published (*51-53*), and we ourselves histologically diagnosed non-specific myositis in some patients in the subacute aftermath of mild or moderate COVID (Fig. S3F). Furthermore, immune-mediated myositis in severe COVID has been well documented by two independent autopsy studies (*29, 30*). We can therefore not exclude that some individuals in our cohort suffered from a self-limiting acute myositis which had resolved by the time of the biopsy.

No SARS-CoV-2 specific RNA could be detected in any of the muscle samples by ultra-sensitive qPCR, strongly arguing against an unresolved infection of skeletal muscle tissues as the cause for the patients’ symptoms.

The mere fact that the number of CD169^+^ macrophages is increased in mildly altered skeletal muscle tissue is remarkable. CD169^+^ macrophages are increased in idiopathic inflammatory myopathies (*54*), indicating a prominent role in type I Interferon-related immune processes (*55*), and recent studies emphasized their highly specific functional programmes and important roles as border-associated cells at blood vessel/parenchymal interfaces (*56, 57*). CD169^+^ macrophages have further been implicated in antiviral defense, being the primary cell infected and able to capture viral particles in the blood and subsequently presenting them to B cells (*58*). As numbers of circulating CD169^+^ monocytes are increased in acute stages of mild COVID-19 (*59*), we hypothesize that they could play a key role at the muscle/capillary interface in our cohort of patients with PCS.

With regards to the fact that most patients from the PCS cohort displayed a selective atrophy of type-2b-fibers - which constitutes a non-specific finding observed in diverse settings leading to disuse or deconditioning of a muscle - we considered the hypothesis that any of our observations could also be the consequence of reduced mobility in these patients due to their exercise intolerance, pain or to coincidental phenomena (lockdowns, stress, mood disorders), instead of being directly related to the infection or to post-infectious mechanisms. We therefore added another historical control cohort to the study, consisting of vastus lateralis samples obtained from patients that displayed a selective atrophy of type-2b-fibers.

On the protein level, mass spectrometry of the vastus lateralis muscle samples revealed an upregulation of basement membrane and other extracellular matrix components only when comparing the PCS samples to the HDC samples but not in comparison to the ones with type-2b-fiber atrophy. Overall, proteome differences were rather subtle, as none of the proteins was increased more than 1.5-fold. This is however not surprising, regarding the fact that histologically little pathology could be noted in the PCS samples. Also, the vastus lateralis muscle samples we used for the HDC cohort came from symptomatic individuals where biopsies were performed for diagnostic purposes, but for which routine histological examination had revealed no morphological abnormalities.

While the muscle samples from the 2BA cohort showed a wider distribution of CBM thickness with some individuals showing similar enlargements as the PCS cohort and higher expressions of CBM-proteins compared to the healthy control group, the mean CBM thickness was still lower in that cohort. We speculate that the 2BA cohort was more heterogeneous than the HDC one, with some of the patients possibly being affected by yet undiagnosed musculoskeletal or systemic diseases that also affected the extracellular matrix. Alternatively, increased matricellular proteins could also be a direct consequence of the selective atrophy of type-2b-fibers, for which little molecular pathomechanistic knowledge exists as of today. The fact that type-2b-fiber atrophy was more pronounced in the 2BA cohort than in the PCS cohort (Fig. 2B) is however an argument against a correlation between type-2b-fiber atrophy and CBM thickness. Moreover, we screened three independent bulkRNA datasets obtained from M. vastus lateralis biopsies performed before and after so-called bed rest studies. Even in this extreme form of immobility with weeklong bedrest, it appears that there is no increased expression of CBM components (collagen IV, laminins, nidogens, heparan sulfate proteoglycans) (*60-62*). Another study, with only a 48h immobilization, revealed a decreased expression of Collagen IV constituents (*63*). These studies however did not include ultrastructural assessment of capillaries, limiting the comparison. Another bed rest study revealed a preserved C/F ratio and increased capillary density due to decreased cross-sectional fiber size (*64*). Of note, macrophage infiltration was associated with the muscular growth phase in a rehabilitation context, rather than the hypotrophic phase of resting (*65*).

On the transcriptional level, unbiased analysis revealed a certain heterogeneity within the PCS cohort, but clustering based on immune cell markers, complement pathway and type I and III interferon related genes allowed a clear separation of the patient’s samples from the controls. PCS samples not only showed an upregulation of immune regulatory genes such as tumor necrosis factor alpha (*TNFA*), but also of genes related to extracellular matrix organization and cell-cell adhesion, while pathways related to oxidative phosphorylation, mitochondria and cell respiration were downregulated. This invites for the speculation that the observed morphological alterations of the capillaries (reduced C/F; thickened CBM) are indeed responsible for metabolic disturbances, possibly explaining the exercise-dependent symptomatology.

From a theoretical, physiological perspective, an increased CBM thickness results in a reduced diffusion of oxygen according to Fick’s Law^ii^. Impaired oxygen delivery to skeletal muscles has been previously described in patients with ME/CFS (*66*), consisting mainly of a reduced peak oxygen uptake during physical activity (*67*) and oxygen therapy has been shown to improve the symptoms (*68*). On the other hand, endothelial damage and capillary pathology have been extensively described in acute as well as in post-acute sequelae of SARS-CoV-2 infection in both human and animal studies (*69-74*). It therefore appears plausible that a capillaropathy contributes or even might be the cause for the described symptoms in a subset of patients with post-COVID syndrome.

Viral infections are well-known triggers for a multitude of autoimmune processes (*75, 76*). Our findings suggest persistent local immune responses in subsets of patients with PCS even one year after initial infection, which in the absence of evidence for an unresolved infection and the presence of autoantibodies in some individuals from our cohort, may point towards immune system dysregulations or an autoreactivity, consistent with multiple observations in patients with acute and post-acute COVID-19 (*77-83*).

To conclude, we hypothesize that acute infection may have caused persistent structural changes of the microvasculature in skeletal muscles in some patients, potentially explaining the exercise-dependent symptoms. The increased presence of CD169^+^ macrophages and the above-mentioned transcriptomic changes at the tissue level approximately one year after infection, together with the absence of SARS-CoV-2-specific RNA suggest that a sustainably dysregulated immune response could be responsible for the microvascular alterations in skeletal muscles of affected patients. Larger studies may allow to identify a “capillaropathy subset” among patients suffering from PCS, ME/CFS or other post-infectious syndromes, thus opening new doors for differential diagnosis and personalized therapies.

The fact that both of our control cohorts consisted of clinically symptomatic patients can be seen both as a strength and a weakness of the study: The weakness lies in the fact that as opposed to non-symptomatic, healthy subjects certain non-specific but pathological changes could be missed on the molecular level in our PCS cohort. The strength is that the observed differences can be attributed to the studied condition with a higher probability, as bystander effects of impaired health status resulting in a less active lifestyle will be leveled out to some degree.

Other limitations are the relatively small cohort of affected individuals, the lack of a control group of asymptomatic patients after a SARS-CoV-2 infection or of a control group with people with similar symptoms but which were not infected with SARS-CoV-2.

## MATERIALS AND METHODS

### Study design

Nine patients who presented to the Charité outpatient clinic because of suspected post-COVID syndrome and two patients that were hospitalized at the Charité were included between June 2020 and November 2021 based on the following criteria:

1. Age >18 years
2. PCR-proven SARS-CoV-2 infection
3. Persistent muscular fatigue and post exertional malaise (PEM) first manifesting after infection with SARS-CoV-2 and lasting for at least 6 months
4. Exclusion of other causes explaining the symptoms listed under (3)
5. Approval for and absence of contraindication for vastus lateralis muscle biopsy

All patients signed informed consent before study inclusion and the study was approved by the Ethics Committee of the Charité - Universitätsmedizin Berlin (EA2/066/20 and EA2/163/17) in accordance with the 1964 Declaration of Helsinki and its later amendments.

Nine of the eleven patients were part of a larger prospective observational study of post-COVID-19 chronic fatigue (*14*). These patients were seen at least once in the outpatient clinic, when a detailed clinical evaluation and neurological examination (muscle strength testing of major muscle groups, handgrip strength test, reflexes and sensory testing, 6-minute-walk-test) was performed and serum samples were obtained. On at least one other occasion, study participants responded to online questionnaires (Bell and Chalder fatigue questionnaires, EQ-5D-5L) (*84, 85*) hosted in a secure REDCap database as previously described (*14*). Proximal lower extremity MRI was performed in these patients on the same day or close to the day of the biopsy. Two other patients (*PCS-7* and *-10*) were included out of an inpatient setting based on the above-mentioned inclusion criteria. After inclusion, one of them (*PCS-7*) was diagnosed with a rheumatoid arthritis and primary biliary cholangitis. Clinical records were consulted for age, sex, preexisting medical conditions, onset and nature of acute and chronic clinical symptoms, laboratory results, therapeutic measures, and complications. All included patients were contacted by telephone beginning of December 2022 for a final follow-up evaluation, assessing the current state of perceived symptoms.

### Vastus lateralis muscle biopsy

Biopsies were performed according to standard procedures as previously described (*86*). In short, after informed consent for the procedure had been granted, open biopsy of the vastus lateralis muscle was performed under local anesthesia (lidocaine 2 %). After circumscribed incision of the skin with removal of a 2 × 5 mm section of the cutis, the muscle was carefully removed. A 15 × 15 mm part of the skeletal muscle, and a 5 × 10 mm part of the muscle fascia were acquired for histopathological assessment. After biopsy procedure at the *Department of Neurosurgery at Charité – Universitätsmedizin Berlin*, muscle tissue, and fascia were processed immediately at the *Department of Neuropathology, Charité – Universitätsmedizin Berlin*.

### Control cohorts

Cryopreserved skeletal muscle specimens obtained from the vastus lateralis muscle were selected based on the following inclusion criteria:

1. Age between 18 and 65 years
2. Vastus lateralis muscle biopsy prior to December 2019
3. Absence of known inflammatory disease, cancer or mitochondriopathy
4. Absence of increased creatinine kinase levels, pathological EMG, corticosteroid or other immunosuppressive therapy
5. For the healthy disease control (HDC) cohort: absence of any histopathological or immunohistochemical abnormality in the excised muscle tissue
6. For the type-2b atrophy control cohort (2BA): presence of a selective atrophy of type-2b-fibers but other than that absence of any histopathological or immunohistochemical abnormality in the excised muscle tissue

These biopsies had been performed for routine diagnostic reasons, and patients had consented to further processing of their samples for scientific purposes. We chose the term “healthy disease control (HDC)” as these patients were clinically diseased (symptoms reported in Extended Data Table 1) justifying a muscle biopsy, but did not show any histological abnormalities. Due to the high prevalence of women in our PCS cohort, we preferentially selected samples of women fulfilling the above-mentioned criteria. However, due to restricted numbers of available samples from women that fulfilled all our inclusion criteria, the HDC group was composed of five men and three women. The type-2b-fiber atrophy control cohort on the other hand consisted of women only. The mean age of the HDC group was 43 years (SD 11; median 45), the one of the 2BA group was 45 years (SD 14; median 45).

### Magnetic Resonance Imaging (MRI)

MRI scans were acquired on a 3 Tesla scanner (MAGNETOM PRISMA®, Siemens, Erlangen, Germany). The subjects were examined in the supine position and feet first using a 28-channel sensitivity encoding torso array coil placed anteriorly. Total scan duration was approximately 35 minutes and included qualitative imaging by axial and coronal T2-weighted turbo spin echo (TSE), axial T1-weighted TSE, axial T1-weighted 3D volumetric interpolated breath-hold examination (VIBE) with Dixon fat suppression and reconstruction of in-/opposed phase, water- and fat-based images, axial 2D Spin-Echo (SE) T2 mapping, details are contained in Extended Data Fig 1c. The field of view (FOV) was at mid-thigh level and anatomic T1w/T2w imaging, T2 maps, and DTI were acquired using the same FOV and geometry. The mean diffusivity (MD), T1 and T2 relaxation times, and muscle quantitative fat fraction (MFF [%]) were evaluated using Visage Imaging Client (Software Release v7.1, Visage Imaging). Manual seeding of regions-of-interest (ROI) at mid-thigh level avoiding areas of fatty infiltration or vascular structures in the biceps femoris (BF), semitendinosus (ST), semimembranosus (SM), and vastus lateralis (VL) muscle was conducted by a radiologist with more than 8 years’ experience in musculoskeletal MRI, blinded to the patients’ clinical data. The MFF was calculated using axial 3D gradient echo-modified two-point Dixon-based MRI with a chemical shift-encoded reconstruction of the water and fat signal as i) SIFAT / (SIFAT + SIWATER) x 100 and reported as mean value of all pixels within the ROI. One patient (*PCS-3*) interrupted the examination before the MD and T2 relaxation times were acquired.

### Virological analysis

Unfixed, cryopreserved muscle samples were used for detection and quantification of SARS-CoV-2 RNA by quantitative reverse transcription–polymerase chain reaction (RT-qPCR). Only samples with at least two positive results were considered positive. Oligonucleotides targeting the leader transcriptional regulatory sequence and a region within the single-guide RNA encoding the SARS-CoV-2 E gene were used to detect single-guide RNA as described previously (*87, 88*).

Anti-SARS-CoV-2 IgG enzyme-linked immunosorbent assays with S1 and NCP domain substrate were performed in available serum samples according to the manufacturer’s instructions (Euroimmun AG®, Lübeck, Germany). In addition, electrochemiluminescence immunoassay (*ECLIA*) antigen tests were performed to detect S- and N-antigens according the manufacturer’s instructions (Elecsys®, Roche, Basel, Switzerland).

### Autoimmunity assays

Antinuclear antibody assays (HEp2 –IFT), myositis-specific autoantibodies (anti–nuclear matrix protein-2 [anti-NXP2], anti–transcriptional intermediary factor 1γ [anti-TIF1γ], anti–melanoma differentiation-associated gene 5 [anti-MDA5], anti–signal recognition particle [anti-SRP], anti-Mi2, anti-isoleucyl-transfer RNA [tRNA] synthetase [anti-OJ], anti–glycyl-tRNA synthetase [anti-EJ], anti–threonyl-tRNA synthetase [anti-PL7], anti–alanyl-tRNA synthetase [anti-PL12], anti-histidyl-tRNA synthetase [anti-Jo1], and anti–small ubiquitin-like modifier-1 activating enzyme [anti-SAE]), and myositis-associated autoantibodies (anti-Ku, anti-PM75, anti-PM100, and anti-Ro52) were performed in available serum samples according to the manufacturer’s instructions (ANA-Mosaik 1A EUROPattern and EUROLINE ANA-Profil 3 (IgG), EUROIMMUN Medizinische Labordiagnostika AG, Lübeck, Germany).

### Histology & Immunohistochemistry

Unfixed biopsy specimens were snap-frozen in a container with isopentane in liquid nitrogen and stored at -80 °C until further workup. Stainings on cryopreserved samples were performed on 7μm thick cryomicrotome sections. Routine histological and enzymological staining (hematoxylin-eosin, Gömöri trichrome, periodic acid–Schiff, ATPases) were carried out according to standard procedures. Immunohistochemical staining was performed on a Benchmark XT autostainer (Ventana Medical Systems), as described previously (*89*).

For quantification of immune cell populations (CD68-, CD169-, CD206-, CD45- and CD8-positive cells, respectively) and semiquantitative scoring (degree of MHC class I & II upregulation and of type-2b-fiber atrophy), ten random fields of vision were examined independently by two experienced morphologists (W.S. and T.A.) at ×400 magnification with an Olympus BX50 microscope (Ocular WH10X-H/22). Positively stained immune-cells were counted manually in 10 high-power fields of vision. Positive staining results with MHC class I and MHC class II were defined as a clear upregulation at the sarcolemma with capillaries and arterioles serving as internal positive controls. The following antibodies were used: MHC class I (DAKO; clone W6/32, 1:100), MHC class II (DAKO; M0775, 1:100) CD45 (DAKO; clone UCHL1, 1:100), CD68 (DAKO; clone EBM11, 1:100), CD8 (DAKO; clone C8/144B, 1:100), NKp46 (R&D Systems; clone MAB1850, 1:100), Siglec-1/CD169 (Novus Biologicals; clone HSn 7D2, 1:200), C5b-9 (DAKO/M777; clone aE11 1:100), CD206 (Abnova clone 5C11; 1:50).

### Transmission Electron Microscopy and capillary morphometry

Electron microscopy was performed as described previously (*88*). In short, after fixation in 2.5% glutaraldehyde in 0.1 M sodium cacodylate buffer muscle samples were incubated with 1% osmium tetroxide in 0.05 M sodium cacodylate and embedded in Renlam resin after dehydration by a graded acetone series. Semithin sections of 500 nm were cut with an ultramicrotome (Ultracut E, Reichert-Jung) and a Histo Jumbo diamond knife (Diatome) and stained with toluidine blue at 80°C. Ultrathin sections of 70nm were cut using the same ultramicrotome and an Ultra 35° diamond knife (Diatome) and stained with uranyl citrate. Standard transmission EM was performed using a Zeiss 906 microscope in conjunction with a 2k CCD camera (TRS).

For each patient, between 20 and 30 randomly selected capillaries from at least two different Renlam resin blocks were photographed at a final magnification of 7000×. Blurry images with unclear basement membrane borders as well as images of abnormally large microvessels or capillaries with very high pericyte coverage were excluded from the analysis. Capillary basement membrane (CBM) thickness was measured at six distinct sites per capillary with *Image J v1*.*53c* (NIH, Bethesda, MD, United States), omitting areas close to profiles of pericyte processes where the CBM is irregular and usually thicker at these sites. For the evaluation of compartmental organization of capillaries, micrographs showing capillary profiles with an aspect ratio (ratio of the smallest to largest diameter) of more than 1.2 were considered too obliquely sectioned and were excluded from morphometric evaluation, as previously recommended (*90*).

Tablet-based image analysis (TBIA) was performed for capillary morphometry, as previously described (*91*). On electron micrographs of capillary profiles, lines were drawn with a digital pen using *ImageJ* around the capillary lumen (blood:EC transition), along the abluminal EC surface (EC:BM transition), along the basement membrane (BM):endomysium transition, and around the PC surface to obtain values for areas and circumferences. Absolute arithmetic values for the lumen radius, and the EC and BM thicknesses were calculated using formulae previously described (*92*). For subjective scoring of capillary alterations, images were interpreted in a blinded fashion, side-by-side by two neuropathologists (W.S. and H.H.G.) and independently by one physiologist (O.B.) with long-standing experience in ultrastructural analysis of skeletal muscle capillaries.

### Morphometry of light micrographs for the evaluation of capillarity

Semithin sections were prepared as described above and per patient at least 10 images at a magnification of 400x were taken with a Keyence BZ-X810. After anonymization of the obtained images, each of these light micrographs was overlaid with a digital counting grid consisting of 10 × 10 test lines, as previously reported (*93*). For calculation of the capillary-to-fiber (C/F) ratio, the number of capillary profiles and that of muscle fibers were counted within the counting grid, taking into account the so-called forbidden line rule. For an estimate of the mean cross-sectional fiber area (MCSFA), the number of test points falling on fiber profiles was divided by the total number of test points and multiplied by the total area of the counting grid (= total area of muscle fiber profiles) divided by the number of muscle fiber profiles that fell within the confines of the grid. The capillary density was determined by dividing the number of capillary profiles that fell within the boundaries of the grid by the total area of muscle fiber profiles.

### Muscle sample preparation for proteomics

Muscle specimens were lysed in 200 µl of 50 mM TEAB (pH 8.5) buffer, 5% SDS, and complete ULTRA protease inhibitor (Roche) using the Bioruptor® (Diagenode) for 10 minutes (30 seconds on, 30 seconds off, 10 cycles) at 4 °C. To ensure complete lysis we conducted an additional sonication step using an ultra-sonic probe (30s, 1s/1s, amplitude 40%) followed by centrifugation at 4°C and 20,000 g for 15 min. Protein concentration of the supernatant was determined by BCA assay according to the manufacturer’s protocol. Disulfide bonds were reduced by addition of 10 mM TCEP at 37°C for 30 min, and free sulfhydryl bonds were alkylated with 15 mM IAA at room temperature (RT) in the dark for 30 min. 100 µg protein of each sample was used for proteolysis using the S-Trap protocol (Protifi) and using a protein to trypsin ratio of 20:1. The incubation time for trypsin was changed to 2 h at 47°C.

All proteolytic digests were checked for complete digestion after desalting by using monolithic column separation (PepSwift monolithic PS-DVB PL-CAP200-PM, Dionex) on an inert Ultimate 3000 HPLC (Dionex, Germering, Germany) by direct injection of 1 μg sample. A binary gradient (solvent A: 0.1% TFA, solvent B: 0.08% TFA, 84% ACN) ranging from 5-12% B in 5 min and then from 12-50% B in 15 min at a flow rate of 2.2 μL/min and at 60°C, was applied. UV traces were acquired at 214 nm (*94*).

### Proteomic analysis of muscle samples

An UltiMate 3000 RSLC nano UHPLC coupled to a QExactive HF mass spectrometer was used for the analysis of all samples, and the total amount of peptides used was always 1 µg. Samples were first transferred to a 75 µm x 2 cm, 100 Å, C18 precolumn at a flow rate of 10 µl/min for 20 minutes followed by separation on the 75 µm x 50 cm, 100 Å, C18 main column with a flow rate of 250 nl/min and a linear gradient composed of solution A (99.9% water, 0.1% formic acid) and solution B (84% acetonitrile, 15.9% water, 0.1% formic acid), with a pure gradient length of 120 minutes (3-45% solution B). The gradient was applied as follows: 3% B for 20 min, 3-35% for 120 min, followed by 3 wash steps, each reaching 95% buffer B for 3 min. After the last wash step, the instrument was equilibrated for 20 min. MS data were acquired in data independent acquisition (DIA) mode using an in-house generated spectral library for the corresponding tissue or body fluid. Each sample was mixed with an appropriate amount of iRT standard (Biognosys). Full MS scans were acquired from 300-1100 m/z at a resolution of 60,000 (Orbitrap) using the polysiloxane ion at 445.12002 m/z as the lock mass. The automatic gain control (AGC) was set to 3E6 and the maximum injection time was set to 20 milliseconds. The full MS scans were followed by 23 DIA windows, each covering a range of 28 m/z with an overlap of 1 m/z, starting at 400 m/z, acquired at a resolution of 30,000 (Orbitrap) with an AGC of 3E6 and an nCE of 27 (CID). For the analysis of samples acquired by nano-LC-MS/MS in DIA mode, the data were entered into Spectronaut software (Biognosys) and analyzed using a library-based search. The library utilized were the in-house created spectral libraries, depending on the tissue or body fluid type. The search and extraction settings were kept as default (BGS Factory settings). Human proteome data from UniProt (www.uniprot.org) with 20,374 entries were selected as the proteome background. For reliable label-free quantification, only proteins with ≥ 2 unique peptides were considered for further analysis. The average normalized abundances were determined using Spectronaut for each protein and used to determine the ratio between the patients’ samples and the corresponding controls. The significance of the abundance changes were calculated with DESeq (*95*).

### Quantitative reverse transcription PCR (qRT-PCR)

Total RNA was extracted from muscle specimens using a trizol-chloroform method as described previously (*89*) and cDNA was synthesized using the High-Capacity cDNA Archive Kit (Applied Biosystems, Foster City, CA, USA). For qPCRs, 10ng of cDNA was used for subsequent analysis, using an Applied Biosystems(tm) QuantStudio(tm) 6 Flex Real-Time PCR System (ThermoFischer, Waltham, MA, USA) with the following running conditions: 95°C for 20 s, 95°C for 1 s, 60°C for 20 s, for 45 cycles (values above 40 cycles were defined as not expressed). All targeted transcripts were run as triplicates. For each of these runs, the reference gene *GAPDH* has been included as an internal control to normalize the relative expression of the targeted transcripts. The qPCR assay identification numbers, TaqMan® Gene Exp Assay from Life Technologies/ ThermoFisher are listed as follows: *GAPDH* Hs02786624_g1, *COL4A1* Hs00266237_m1, *COL4A2* Hs05006309_m1, *NID1* Hs00915875_m1, *NID2* Hs00201233_m1, *HSPG2* Hs01078536_m1. The ΔCT of HDCs was subtracted from the ΔCT of COVID patients muscles to determine the differences (ΔΔCT) and fold change (2^-ΔΔCT) of gene expression.

### Bulk RNA-sequencing

RNA was isolated using Trizol (ThermoFisher) and the DirectZol kit (Zymo) according to manufacturer’s instruction. Poly-A RNA sequencing libraries were then prepared using the NEBNext Ultra II Directional RNA Library Prep Kit (NEB), and sequenced to a depth of 20-30 million paired-end reads on a NovaSeq 6000 device (Illumina) with 2&109 sequencing length. Since the samples were sequenced together with samples containing high amounts of SARS-CoV-2 RNA on the same Novaseq sequencing lane, there were some reads aligning to the SARS-CoV-2 genome due to index hopping on the flowcell, i.e. misassignement of samples with high SARS-CoV-2 amount to the muscle samples. Due to this effect, quantification of SARS-CoV-2 RNA from sequencing is less reliable than from RT-qPCR.

### RNA sequencing analysis

Due to data protection restrictions, raw sequencing reads cannot be made readily available, however a read count table is provided as supplementary data. Raw sequencing reads were aligned to version hg19 of the human genome using hisat2 (*96*) with standard parameters. Read counts were quantified based on the hg19 RefSeq annotation using quasR (*97*). Differential expression values were calculated using edgeR (*98*). Further analysis was done and plots were generated using the R packages PCAtools [PCAtools: Blighe K, Lun A (2022). PCAtools: PCAtools: Everything Principal Components Analysis. R package version 2.10.0, https://github.com/kevinblighe/PCAtools], ComplexHeatmap [ComplexHeatmap: Gu Z (2022). “Complex Heatmap Visualization.” iMeta. doi: 10.1002/imt2.43.], as well as packages from the Tidyverse [https://doi.org/10.21105/joss.01686]. All code is available on github.com/LandthalerLab.

### GO Enrichement analysis and Metascape pathway analysis of proteomic and transcriptomic data

For GO Enrichement analysis (*99-101*) of the proteomic data sets, significantly regulated proteins (p-value ≤0.01) with either positive or negative regulation (calculated fold change of ≥1.2or ≤0.8) were included. For pathway analysis of the transcriptomic data sets, significantly regulated genes (p-value ≤0.01) with either positive or negative regulation (calculated log fold change of ≥0.8 or ≤ -0.8) were included using Metascape (https://metascape.org) (*102*).

### Statistical analysis

Statistical analyses were performed and graphs created with *GraphPad Prism 9* (GraphPad Software) and *R* Software (A language and environment for statistical computing. R Foundation for Statistical Computing, Vienna, Austria; https://www.R-project.org/).

Normality testing was performed (D’Agostino & Pearson, Anderson-Darling, Shapiro-Wilk and Kolmogorov-Smirnov tests) and a Gaussian distribution was confirmed for the following parameters: immune cell quantification (CD68, CD169, CD206, CD45, CD8), capillary-to-fiber-ratio, MCFSA, capillary density, CBM thickness.

One way Anova with Tukey’s multiple comparison test were used to compare differences between the three cohorts. Data are presented as counts, percentages or means (SDs). Values were considered significant at *p* ≤0.05. Heat map and dot plot visualizations were created with *R* Software and tables with *Excel 2016* (Microsoft).

## Data Availability

All data produced in the present study are available upon reasonable request to the authors.

## Acknowledgments

We thank Cordula zum Bruch, Silvia Stefaniak and Petra Matylewski, Department of Neuropathology, Charité Universitätsmedizin, for their excellent technical assistance. They were not compensated for their contributions.

## Author contributions

Writing – original draft, data analysis and visualization, TEM of capillaries: TA

Clinical study design: CS, FP, JBS.

Clinical data acquisition: FL, JBS, LMA, CK, RR, NLD

Muscle biopsies: NLD, AS, PV

Interpretation of histopathology and ultrastructural analysis: WS, HHG, TA

Capillary morphometry (TBIA) and quantification of capillaries: OB

Virology (anti-SARS-CoV-2 antibodies, muscle PCRs): JS, VC

Proteomics data acquisition: AH, AR

Bulk RNA sequencing and analysis: EW, LGTA, ML

qPCR: CP, AF, CD

Writing – review & editing: HHG, HR, FLH, CS, WS

## Data and materials availability

Any data not published within the article or at indicated online locations, can be accessed by any qualified investigator upon demand to the corresponding author.

## Figures

**Table S1.**

| ID | acute COVID symptoms (<14 days) | chronic symptoms (>6 Months) | comorbidities | ME-CFS (CCC) | immuno-suppressive treatment | relevant lab findings | heart MRI |
| --- | --- | --- | --- | --- | --- | --- | --- |
| PCS-1 | loss of taste and smell | Muscular weakness, fatigue, PEM, dysbalance | recurring sinusitis | no | none |  |  |
| PCS-2 | loss of taste and smell, dyspnea, fatigue, arthralgia, cough, rhinitis, hearing impairment, gait disturbance, myalgia, sore throat, cognitive impairment, sleep disturbance, tracheobronchitis | fatigue, PEM, cognitive impairment (memory), dyspnea, burning pain with focus on joints, distal arms and legs | Lyme disease, facial paralysis, hypothyroidism | yes | Prednisolon |  |  |
| PCS-3 | fatigue, dyspnea, headaches, vertigo/dizziness, cough, chest pain/tightness, vomiting/nausea, numbness (legs, body) | shivering, pain (NRS 3), cramps, exercise intolerance, PEM | Asthma, endometriosis, hypothyroidism, neurodermatitis | no | none |  |  |
| PCS-4 | loss of taste and smell, fatigue, tachycardia, body aches, chest pain, back pain, fatigue, subfebrile temperature, rhinitis, sweating | fatigue, PEM, myalgia |  | yes | none | MBL-deficiency, ANA 1:1280, IgG3 subclass deficiency | 05/2020: apical pericarditis<br>02/2021: normal appearance |
| PCS-5 | loss of taste and smell, dyspnea, cough, fever, vertigo, myalgia, body aches | muscular weakness, myalgia, pronounced on arms, PEM | Arterial hypertension, multiple torn ligaments (shoulder, hand, knee, feet) | no | none |  | diffuse fibrotic changes |
| PCS-6 | loss of taste and smell, fatigue, mild dyspnea, chest pain/tightness, fever, sore throat, rhinitis, headaches, chills, night sweat | muscular weakness (full body), PEM | POTS | no | none |  |  |
| PCS-7 | fatigue, myalgia, arthralgia, joint swelling | myalgia, PEM, paresthesia, distal paresis arms, unclassified tremor, hair loss, fatigue, cognitive impairment (Concentration, memory) | Raynaud's phenomenon, primary biliary cholangitis, Polyarthritits | no | Prednisolon; azathioprin; methotrexat | anti-AMA-M2 positive |  |
| PCS-8 | headaches, cognitive impairment | Fatigue, myalgia, PEM, cognitive impairment (concentration), sleep disturbance | Arterial hypertension | no | none |  |  |
| PCS-9 | fatigue, headaches, myalgia/body aches, arthralgia, fever, rhinitis, cough, sore throat | Fatigue, Myalgia, PEM, cognitive impairment (concentration, memory, speech), sleep disturbances, headaches, arthralgias |  | yes | none | MBL-deficiency |  |
| PCS-10 | loss of taste and smell, headaches, body aches, arthralgia, myalgia, cognitive impairment | Myalgia, PEM, fatigue, hyposmia, sleep disturbances, cognitive impairment, headaches |  | yes | none |  |  |
| PCS-11 | Dyspnea requiring non-invasive highflow ventilation for 4 days, fever, cough | Persisting Dyspnea (NHYA II), PEM, exercise intolerance, fatigue, headaches, myalgia, arthralgia, heart palpitations, cognitive impairment (memory, concentration, lacking words), sleep disturbance | Class III obesity | yes | none |  | dilated cardiomyopathy |
| HDC-1 | / | Proximal muscular weakness |  | / | none |  | / |
| HDC-2 | / | Myalgia and muscular weakness legs, intermittend muscle spasms/stiffness | WPW-Syndrome; Tachycardia, aortic aneurysm; hypercholesterolemia | / | none |  | / |
| HDC-3 | / | Unclear movement disorder. Suspected mitochondriopathy was ruled out histopathologically. | tobacco consumption, unclassified tremor | / | none |  | / |
| HDC-4 | / | Muscle hypertrophy right leg of unknown origin; suspected hereditary muscle disease was ruled out |  | / | none |  | / |
| HDC-5 | / | Proximally pronounced progressive muscular weakness and myalgia | Depression, Neurodermatitis, Spondylarthritits | / | none |  | / |
| HDC-6 | / | Exercise intolerance, Post-exertional malaise; muscle cramps and stiffness |  | / | none |  | / |
| HDC-7 | / | General muscular weakness; unilateral ptosis | Myocarditis (in teenage years) | / | none | ANA 1:160; anti-SOX-1 antibodies | / |
| HDC-8 | / | Proximal myalgia and muscular weakness |  | / | none |  | / |
| 2BA-1 | / | Focal epilepsy, progressive global cerebral atrophy. Suspected mitochondriopathy was ruled out histopathologically. |  | / | none | ANA 1:160 | / |
| 2BA-2 | / | Proximal muscular weakness | Arterial hypertension, smoking | / | none |  | / |
| 2BA-3 | / | Exercise-dependent myalgia |  | / | none |  | / |
| 2BA-4 | / | Exercise-dependent muscle weakness. Suspected mitochondriopathy was ruled out histopathologically. | arterial hypertension, hypercholesterolemia, psoriatic arthritis, epilepsy | / | none |  | / |
| 2BA-5 | / | Muscular weakness (arms and legs) | arterial hypertension, migraine, | / | none |  | / |
| 2BA-6 | / | proximal muscular weakness (legs); myalgia |  | / | none |  | / |
| 2BA-7 | / | Exercise-dependent proximal myalgia (legs) | arterial hypertension | / | none |  | / |
| 2BA-8 | / | Myalgia and proximal weakness (legs) | tobacco consumption, peripheral artery disease, aortic aneurysm | / | none |  | / |

**Fig. S1.**
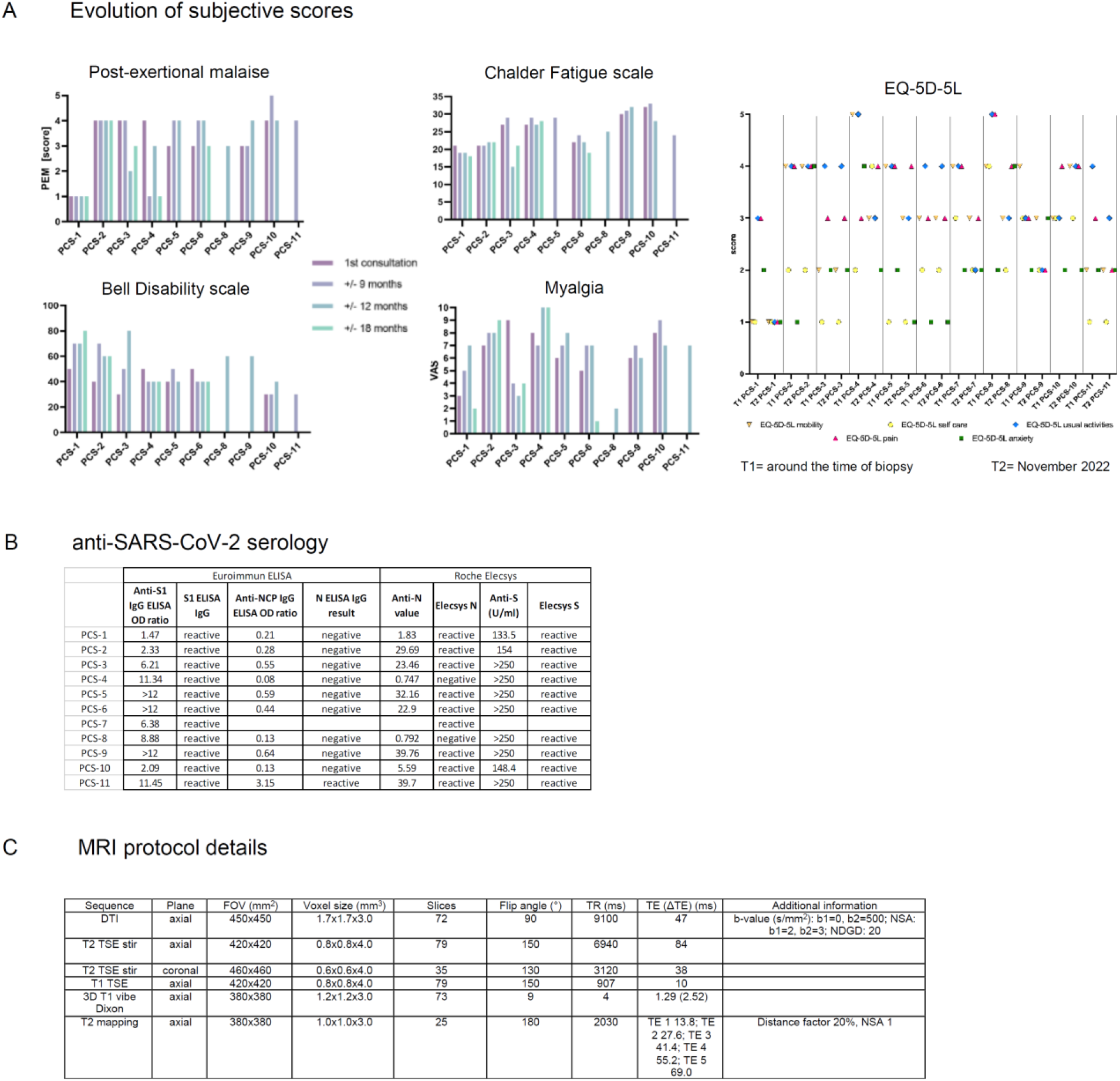
Additional clinical data. (**A**) Evolution of subjective scores at indicated timepoints. For two patients (*PCS-8* and *PCS-11*) indicated scores from only one timepoint were available, and for one patient (*PCS-7*) none was available. (**B**) Detailed results of the serological testing for anti-SARS-CoV-2 N and S IgG. Data were incomplete for *PCS-7*. (**C**) Technical acquisition details of the lower leg MRIs.

**Fig. S2.**
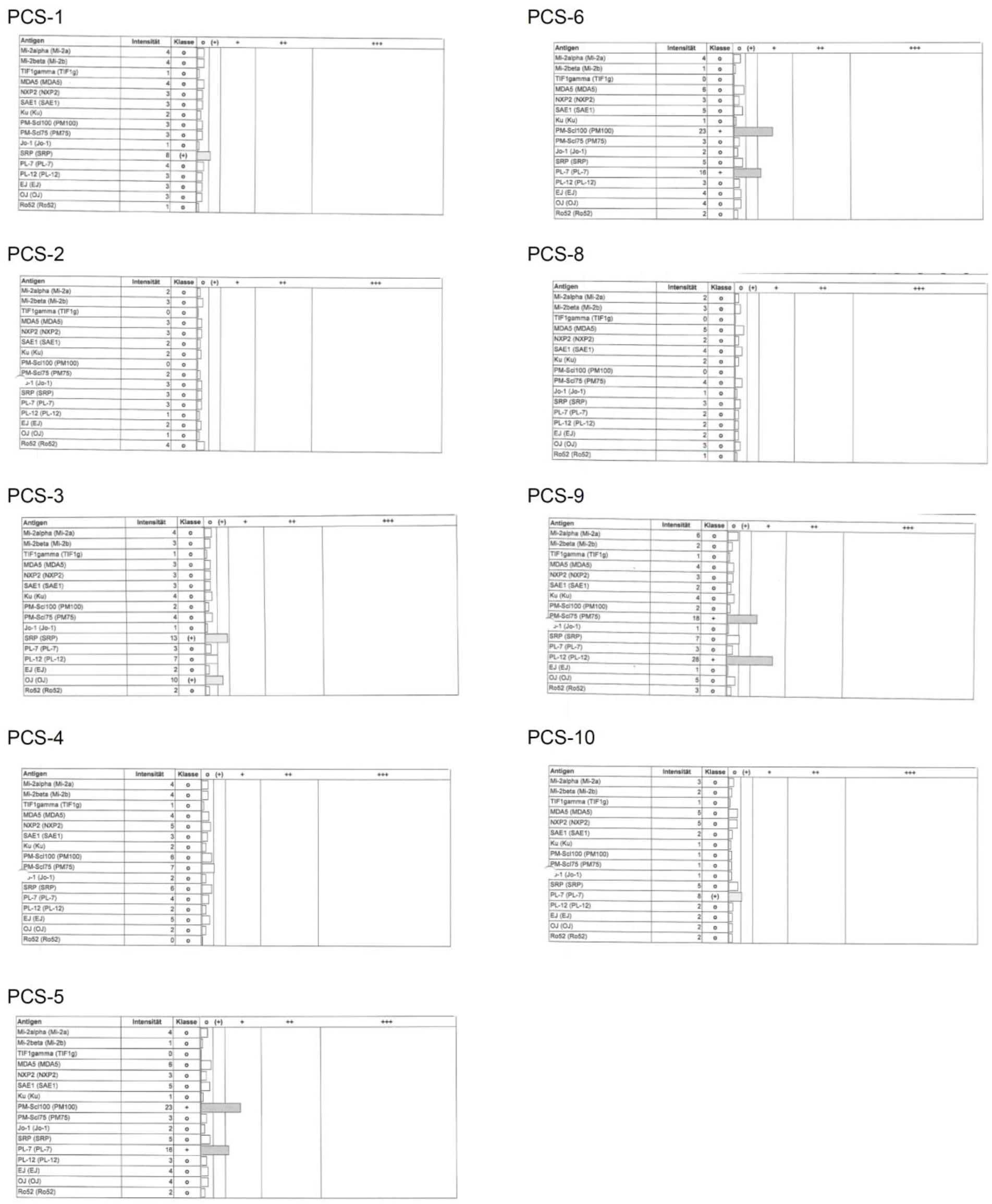
Myositis-related and –associated autoantibody screening. Individual test results of myositis-related and –associated autoantibody screening. No serum was available for this analysis from two patients (*PCS-7* and *PCS-11*).

**Fig. S3.**
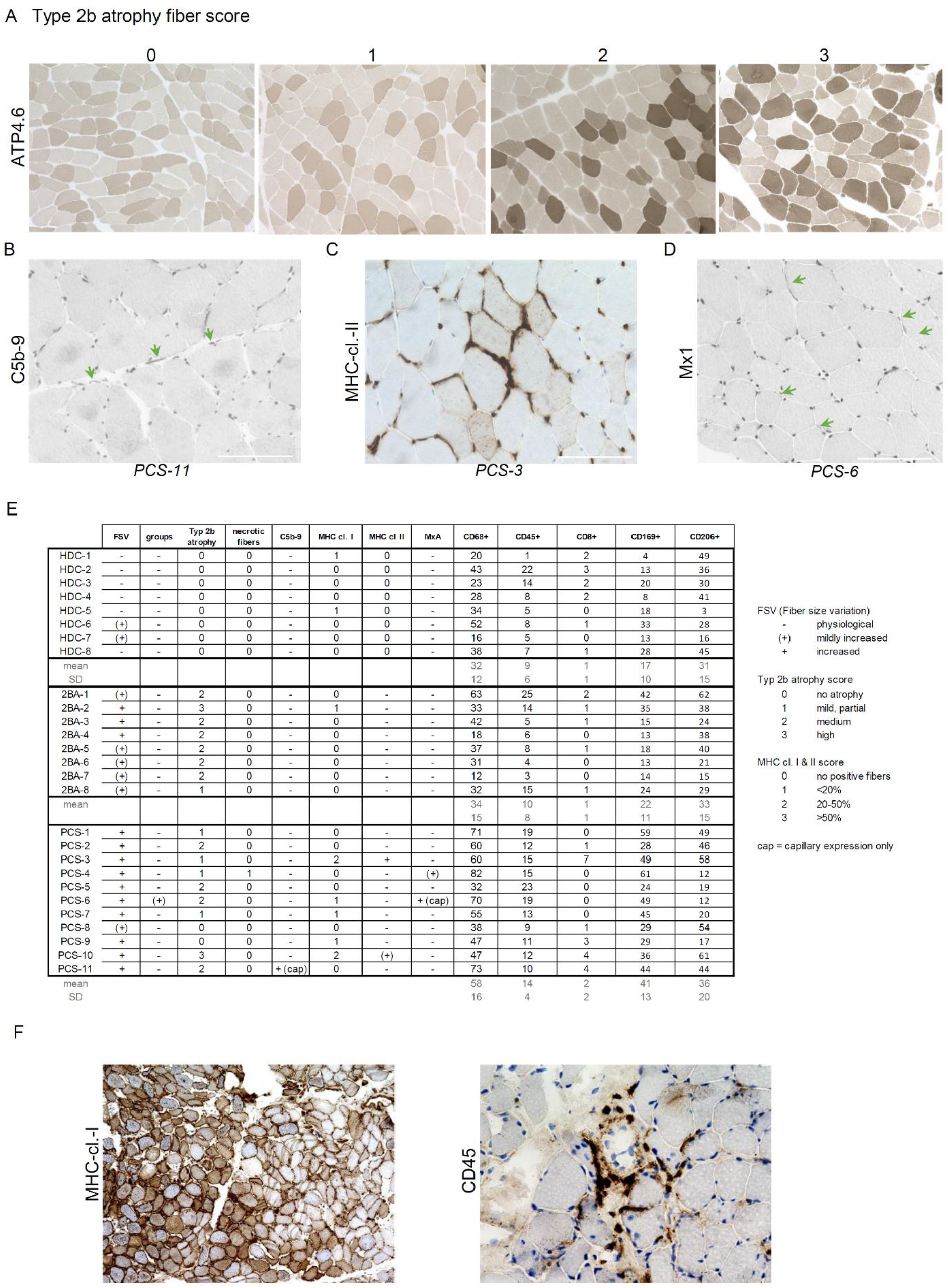
Additional histology. **(A**) ATP4.6 enzymatic reaction of representative samples with different degrees (=scores) of type-2b-fiber atrophy. 200x magnification. **(B**) Immunohistochemistry showing C5b-9^+^ capillaries (green arrows). **(C**) Immunohistochemistry showing sarcolemmal MHC-cl.-II expression. **(D**) Immunohistochemistry showing Mx1^+^ capillaries (green arrows). **(E**) Detailed histopathological findings of all analyzed samples. **(F**) Immunohistochemistry showing post-COVID myositis (CD45^+^ cell infiltrates and sarcolemmal MHC-cl.-I upregulation) in a patient which was not included in the PCS cohort. *Abbreviations: ATP4*.*6= Adenosine 5’-TriPhosphatase at pH 4*.*6; MHC cl. II= major histocompatibility complex class II*.

**Fig. S4.**
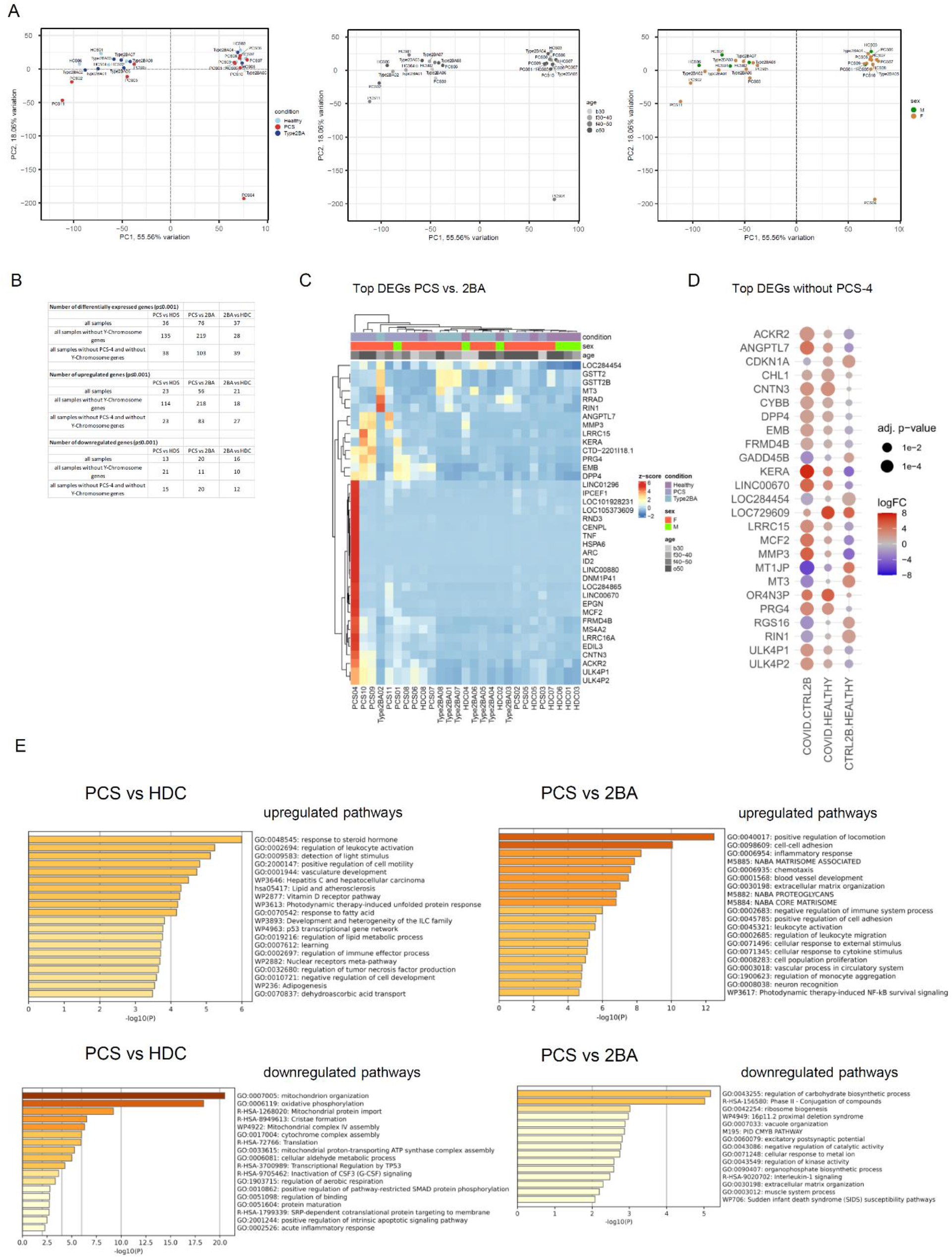
Additional transcriptomic analysis. (**A**) Principal component analysis of the bulk RNA dataset showing absence of clustering by age and sex. (**B**) Summary table of the number of differentially expressed genes (DEGs) considering all samples and all genes, all samples but not Y-Chromosome genes, and all samples except *PCS-4* without Y-Chromosome genes respectively. (**C**) Clustered heat map visualization of top differentially expressed genes comparing the PCS cohort to the HDC and 2BA cohorts. (**D**) Dot plot visualization of top differentially abundant proteins comparing all samples from the PCS cohort minus *PCS-4* to the HDC and 2BA cohorts respectively. Dot size is proportional to the adjusted p-value. Dot colour indicates logFC. (**E**) Metascape pathway analysis of the significantly regulated genes (p-value ≤0.01) with either positive or negative regulation (calculated log fold change of ≥0.8 or ≤ -0.8) comparing the PCS cohort to the HDC or 2BA cohort respectively.

**Fig. S5.**
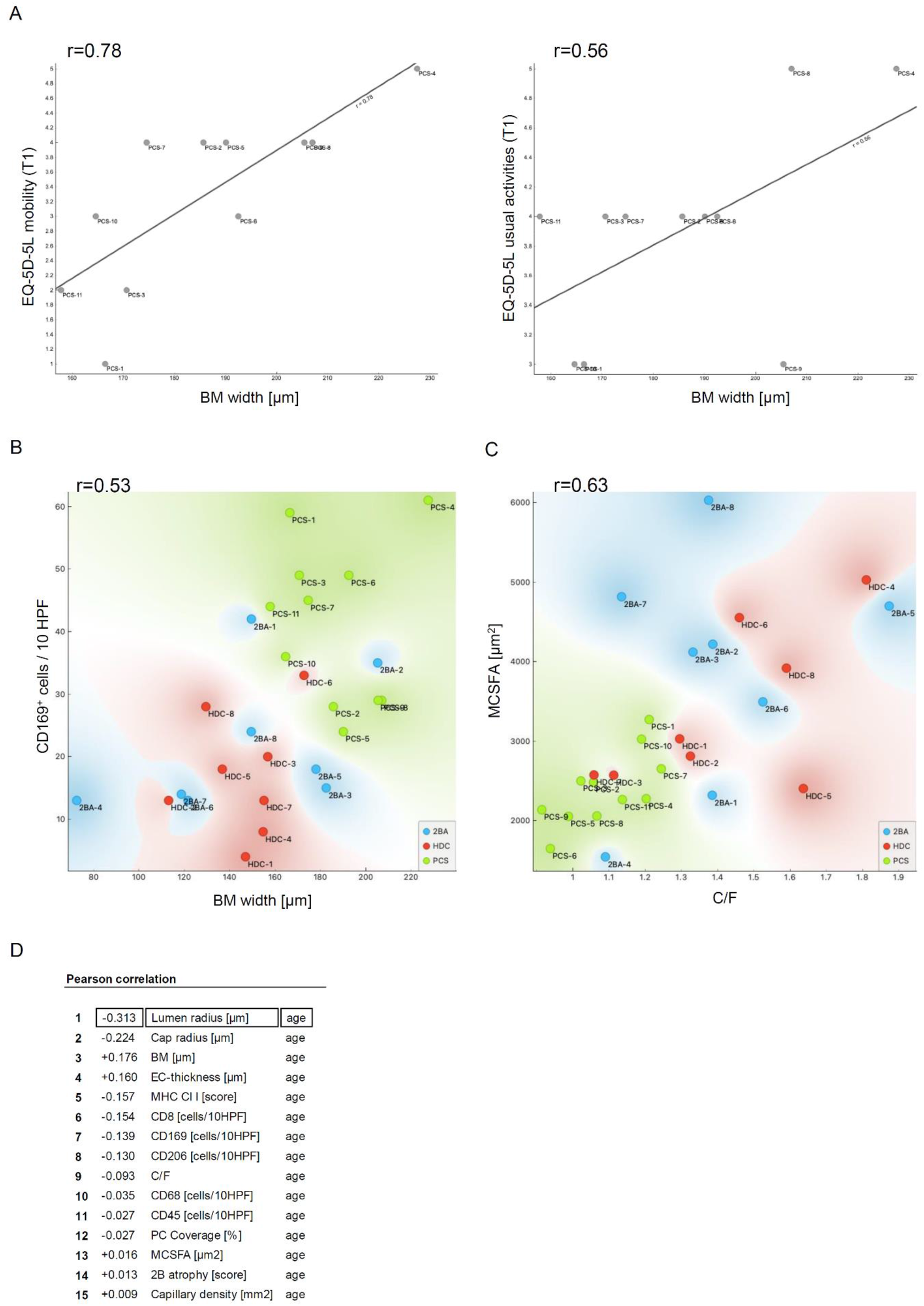
Correlations. **(A)** Correlation of CBM thickness with the EQ-5D-5L mobility score (Pearson’s r 0.78) and the EQ-5D-5L usual activities score around the time of biopsy (Pearson’s r 0.56). **(B)** Correlation of CBM thickness with the number of CD169^+^ macrophages (Pearson’s r 0.53). **(C)** Correlation between the mean cross sectional fiber size area (MCSFA) and the capillary-to-fiber ratio (Pearson’s r 0.63). **(D)** Pearson’s correlation of age and relevant quantitative parameters.

## Footnotes

i Bell, D. S. *The doctor*’*s guide to chronic fatigue syndrome: Understanding, Treating and Living with CFIDS. Boston: Da Capo Lifelong Books* (1995).

ii 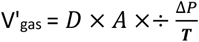 [V’_gas_= *Degree of diffusion of the gas across a permeable membrane*, D = *Diffusion coefficient of the gas and the membrane*, A = *Surface of the membrane*, ΔP = *Difference in partial pressure of the gas across the membrane*, T = thickness of the membrane].

